# An Integrated Multi-Method Framework for Gender-Based Violence Research: A Synthetic Data Demonstration Using Kenya Demographic and Health Survey Parameters

**DOI:** 10.64898/2025.12.15.25342247

**Authors:** Grold Otieno Mboya

**Affiliations:** School of Health Sciences, Jaramogi Oginga Odinga University of Science and Technology (JOOUST), Bondo, Kenya

**Keywords:** Gender-Based Violence, Machine Learning, Causal Inference, Kenya, Epidemiology, Simulation, Random Forest

## Abstract

**Background:** Current research on Gender-Based Violence (GBV) typically separates predictive machine learning and causal inference into distinct analytical silos. Yet, grasping the multi-level determinants of violence requires an approach that can both identify high-value predictors and disentangle their causal mechanisms.

**Methods:** This study develops and demonstrates an integrated five-phase analytical framework for GBV research, applying sequential methods to a single synthetic dataset (N = 3,000) parameterized to reflect the prevalence patterns and risk factor distributions of the 2022 Kenya Demographic and Health Survey (KDHS). The framework was applied across five stages: descriptive epidemiology, Random Forest variable selection, logistic regression for adjusted associations, mediation analysis, and evidence synthesis.

**Results:** The Random Forest model achieved 74.6% accuracy (AUC = 0.711; sensitivity = 37.3%; specificity = 89.8%), recovering partner alcohol use, childhood trauma, and marital conflict as the top-ranked predictors. Importantly, this accuracy offers only a modest gain over the null classifier baseline of 73.1%. Multivariable logistic regression yielded stable adjusted effect estimates for partner alcohol use (aOR = 6.60) and childhood trauma (aOR = 1.99). Mediation analysis tested three theoretically informed indirect pathways; however, none of the hypothesized indirect effects reached statistical significance (ACME p > 0.05 for all pathways), indicating that the specified risk factors operated primarily through direct rather than mediated routes in this simulation.

**Conclusions:** This proof-of-concept demonstration shows that the five-phase framework can be coherently applied to synthetic GBV data parameterized from KDHS estimates. The framework’s multi-level logistic regression approach produced meaningfully better model fit than single-level alternatives (ΔAIC = 67.5). Future research should apply this framework to real-world longitudinal data to rigorously evaluate its performance characteristics and causal inference capacity.

## Background

Gender-based violence (GBV) constitutes a severe violation of human rights and a critical public health challenge with profound implications for women’s physical, mental, and reproductive health. The World Health Organization (WHO) estimates that approximately one in three women worldwide have experienced physical or sexual violence by an intimate partner or non-partner sexual violence in their lifetime [1]. The burden is disproportionately high in sub-Saharan Africa, where lifetime prevalence estimates in some regions exceed 40% [2]. In Kenya, despite legislative frameworks such as the Sexual Offences Act (2006), the prevalence of intimate partner violence (IPV) remains staggeringly high. Data from the Kenya Demographic and Health Survey (KDHS) indicate that 41% of ever-married women have experienced physical, sexual, or emotional violence by a spouse [3].

Beyond immediate physical injury, GBV is strongly associated with adverse long-term outcomes, including depression, post-traumatic stress disorder (PTSD), and suicidal ideation [4, 5]. It is also a known risk factor for HIV acquisition and unintended pregnancies [6].

Recognizing these complex consequences, researchers have increasingly adopted the ecological framework proposed by Heise [7], which posits that violence results from the interplay of factors across four nested levels: individual, relationship, community, and societal.

However, while the ecological framework is widely invoked conceptually, its empirical application often faces significant methodological limitations. Current epidemiological research on GBV is characterized by a “methodological singularity.” Most studies rely exclusively on traditional multivariable regression models [8, 9]. While effective for estimating adjusted associations and controlling for confounding, these models often assume linearity and struggle to handle the high-dimensional, non-linear interactions inherent in ecological systems. Conversely, machine learning (ML) approaches, such as Random Forests, offer superior predictive accuracy and can naturally handle complex interactions [10], but they are frequently criticized in public health for lacking interpretability and causal validity [11].

This dichotomy presents a critical gap in GBV research: studies either predict risk well but fail to explain mechanisms (ML), or estimate average associations well but miss complex, non-linear patterns (traditional regression). Furthermore, while risk factors are frequently identified, the causal pathways—the *mechanisms* through which distal factors like childhood trauma influence proximal outcomes like IPV—remain under-investigated through formal mediation analysis [12].

To address these gaps, this study proposes and demonstrates an integrated analytical framework. By combining the predictive power of machine learning with formal regression-based inference, this research moves beyond identifying *who* is at risk to understanding *why* and *how* that risk manifests. Specifically, this study utilizes a simulation approach based on KDHS structural parameters to: (1) demonstrate how machine learning can inform variable prioritization for regression models; (2) empirically test the explanatory power of the multi-level ecological framework against single-level models; and (3) test mechanistic pathways through mediation analysis. This methodological integration offers a replicable blueprint for future research, ensuring that GBV investigations are grounded in both predictive and associational evidence.

## Theoretical framework

### The Ecological Model

This study adopts Heise’s integrated ecological framework [7] as its organizing theoretical structure. The model posits that GBV risk is not determined by a single factor but results from the interplay of variables operating across four nested levels:

- **Individual (Ontogenic) Level:** This level encompasses the personal history and biological characteristics an individual brings to their relationships. Key factors include age, education, socioeconomic status, and developmental experiences such as childhood trauma or exposure to family violence.
- **Relationship (Microsystem) Level:** The immediate context in which abuse occurs. This includes partner behaviors (e.g., alcohol use), relationship dynamics (e.g., communication quality, conflict resolution), and power differentials regarding decision-making and economic control.
- **Community (Exosystem) Level:** The formal and informal social structures that embed the individual and relationship. Relevant factors include neighborhood poverty concentration, community norms regarding gender roles, and institutional factors such as the availability of support services.
- **Societal (Macrosystem) Level:** The broad cultural values and structural features that permeate the lower levels, including patriarchal norms, legal frameworks, and macroeconomic systems.

### Operationalization and Conceptual Model

For this simulation study, the ecological framework guided both the generation of the synthetic dataset and the specification of the analytical models. While societal-level factors (Macrosystem) were held constant to represent the single national context of Kenya, the analysis explicitly models interactions across the remaining three levels.

The conceptual model tests five specific hypotheses (H):

- **H1:** GBV risk increases with exposure to individual vulnerability factors, specifically childhood trauma and lower socioeconomic status.
- **H2:** Relationship-level factors, particularly partner alcohol use and power imbalances, demonstrate the strongest direct associations with GBV.
- **H3:** Community-level factors (e.g., patriarchal norms) influence GBV both directly and indirectly by modifying relationship dynamics.
- **H4:** A comprehensive ecological model including all three levels provides superior statistical fit compared to models restricted to single levels.
- **H5:** Risk factors operate through specific causal pathways, notably:

- Partner alcohol use → Marital conflict → GBV
- *Childhood trauma → Mental health → GBV*

### Analytical Strategy

To test these hypotheses, the study employs a multi-method approach designed to overcome the limitations of single-method studies. The analysis proceeds through five complementary phases:

1. **Descriptive Epidemiology:** Establishes prevalence and identifies initial bivariate associations.
2. **Machine Learning (Random Forest):** Identifies high-priority predictors without assuming linearity, allowing for the detection of complex interactions often missed by traditional models.
3. **Multivariable Logistic Regression:** Estimates adjusted odds ratios for independent risk factors to ensure interpretability.
4. **Mediation Analysis:** Formally tests the causal pathways (H5) to identify mechanisms of action.
5. **Synthesis:** Integrates findings across methods to isolate convergent risk factors that show robustness regardless of the analytical technique used.

## Methods

### Study Design and Data Generation

This study employed a cross-sectional simulation design to develop and validate a multi-phase analytical framework for gender-based violence (GBV) research. Unlike empirical studies that rely on pre-existing survey data, this research utilized a synthetic dataset (N = 3,000) explicitly parameterized to reflect the structural properties, demographic distributions, and risk factor prevalences of the 2022 Kenya Demographic and Health Survey (KDHS).

Data simulation was conducted in the R statistical environment (version 4.5.2) using the *simstudy* package. This approach provided full control over the data-generating process (DGP), allowing for the “ground truth” of causal relationships to be known a priori. The simulation specified a hierarchical structure comprising 100 community clusters to mirror the complex survey design typical of demographic health surveys. Risk factor effect sizes were pre-specified based on parameters derived from the KDHS and existing meta-analyses (e.g., setting the odds ratio for partner alcohol use at approximately 6.0).

### Study Population and Sample Size

The simulated cohort represented women of reproductive age (15–49 years) who are or have been in an intimate relationship. The sample size was calculated to ensure adequate statistical power across all analytical phases. Using the standard formula for prevalence estimation:

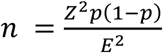

Where Z = 1.96, p = 0.27 (anticipated prevalence), and E = 0.02 (margin of error), the required sample was approximately 1,893 participants. The final sample of N = 3,000 provided a precision of ±1.6% and exceeded requirements for multivariable modeling. With an expected 810 outcome events, the events-per-variable (EPV) ratio was 67.5, well above the recommended minimum of 10 for logistic regression stability. Additionally, this sample size ensured a sufficient 70/30 split (2,100 training / 900 testing) for the machine learning phase.

### Variables and Measures

The primary outcome variable, **Gender-Based Violence (GBV)**, was defined as a binary indicator (Yes/No) representing experience of physical, sexual, psychological, or economic violence by an intimate partner in the past 12 months.

Predictor variables were operationalized across three ecological levels:

- **Individual Level:** Age, educational attainment, wealth quintile, area of residence (urban/rural), history of childhood trauma, and mental health status.
- **Relationship Level:** Partner alcohol use, marital conflict frequency, communication quality, and decision-making power.
- **Community Level:** A community patriarchy index, strength of legal implementation, and geographic distance to support services.

### Statistical Analysis Plan

The analytical framework was executed in five sequential phases.

#### Phase 1: Descriptive Epidemiology

The initial phase involved descriptive analysis to validate the simulated data structure against known KDHS patterns. Continuous variables were summarized using means and standard deviations, while categorical variables were reported as frequencies and percentages. Prevalence estimates for GBV were calculated with Wilson 95% confidence intervals (CIs).

#### Phase 2: Predictive Modeling (Machine Learning)

To identify high-priority predictors without imposing linear assumptions, a **Random Forest** classification model was trained using 500 decision trees. The dataset was split into training (70%) and testing (30%) sets. Variable importance was assessed using the Mean Decrease in Accuracy (MDA) metric, computed via permutation of each predictor in turn. Model performance was evaluated on the test set using the Area Under the Receiver Operating Characteristic Curve (AUC-ROC), overall accuracy, sensitivity, specificity, and F1 score.

#### Phase 3: Causal Inference (Multivariable Regression)

To estimate independent risk factors and test the ecological hypothesis, nested multivariable logistic regression models were fitted sequentially:

1. **Model 1:** Individual factors only.
2. **Model 2:** Individual + Relationship factors.
3. **Model 3 (Full Ecological Model):** Individual + Relationship + Community factors.

Model fit was compared using the Akaike Information Criterion (AIC). Adjusted Odds Ratios (aOR) with 95% CIs were reported for the final model.

#### Phase 4: Causal Mediation Analysis

Mediation analysis was employed to test three theoretically informed pathways identified in the ecological framework:

1. Partner Alcohol → Marital Conflict → GBV
2. Childhood Trauma → Mental Health → GBV
3. Community Patriarchy → Decision Power → GBV

The analysis utilized the potential outcomes framework [13] to decompose total effects into the **Average Causal Mediation Effect (ACME)** and the **Average Direct Effect (ADE)**. For each pathway, a mediator model (M ∼ X) and an outcome model (Y ∼ X + M + Covariates) were fitted.

Significance was determined using 1,000 nonparametric bootstrap samples. Mediation was classified as:

- **Full Mediation:** Significant ACME, non-significant ADE.
- **Partial Mediation:** Significant ACME and ADE.
- **No Mediation:** Non-significant ACME.

Sensitivity analysis was conducted to assess the robustness of findings to the sequential ignorability assumption, calculating the rho (ρ) parameter at which the ACME would become non-significant. All mediation analyses were performed using the *mediation* package in R.

### Phase 5: Sensitivity Analysis

To assess the stability of the framework’s variable selection and estimation across varying conditions, a sensitivity analysis was conducted by re-running the full analytical pipeline at three alternative sample sizes: N = 500, N = 1,000, and N = 5,000, using the same data-generating process and random seed structure. For each scenario, the Random Forest variable importance rankings and the adjusted logistic regression odds ratios for the three primary exposures (partner alcohol use, childhood trauma, and marital conflict) were recorded and compared against the main N = 3,000 results. This analysis evaluated whether the framework’s conclusions are robust to sample size variation and whether smaller samples produce meaningfully different variable prioritization or unstable effect estimates.

### Ethics Statement

This study utilized purely synthetic data generated through computational simulation methods. The simulation parameters were informed by aggregate, publicly available findings from existing literature and demographic surveys. As this research did not involve the recruitment of human participants or the use of identifiable private information, institutional review board (IRB) or ethical approval was not required.

## Results

### Study Sample and Prevalence

The simulated cohort comprised 3,000 women aged 15–49 years, structured to reflect the demographic distribution of the 2022 Kenya Demographic and Health Survey. The overall prevalence of Gender-Based Violence (GBV) in the sample was 26.9% (95% CI: 25.3–28.7%), corresponding to 808 cases.

Table 1 presents the prevalence stratified by key demographic characteristics. Rates were relatively consistent across age groups but varied by wealth and education. Women in the lowest wealth quintile reported a higher prevalence (29.6%) compared to those in the highest quintile (23.2%). Similarly, women with only primary education showed the highest prevalence (30.0%), whereas those with higher education had the lowest (23.2%)

**Table 1.**
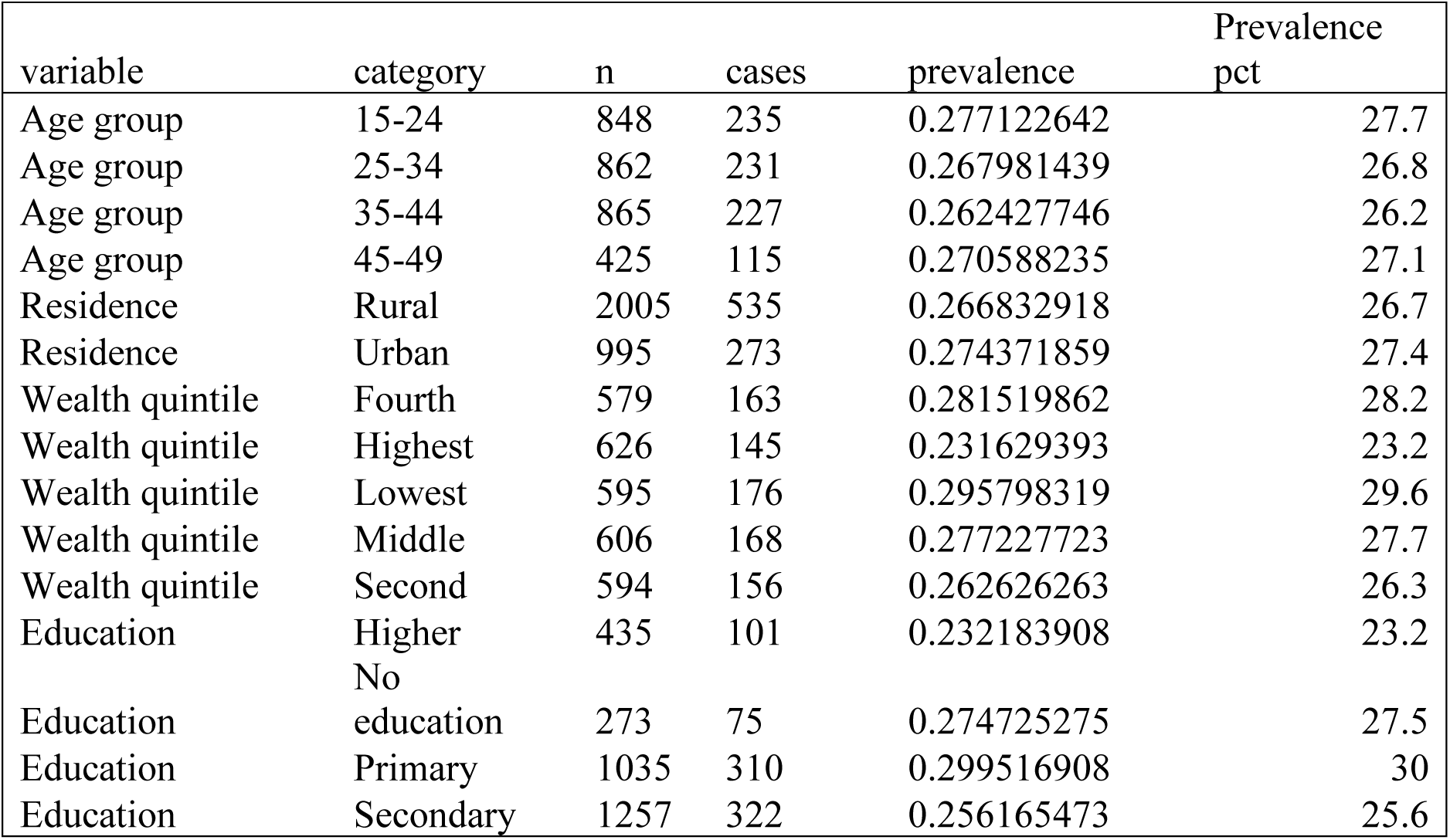
GBV Prevalence by Demographic Subgroups.

### Bivariate Associations

Initial bivariate analyses indicated that all continuous and binary predictors showed associations with the outcome. Partner alcohol use demonstrated the strongest correlation with Gender-Based Violence (GBV) (φ = 0.385, p < 0.001), followed by marital conflict (r = 0.111, p < 0.001) and childhood trauma (φ = 0.105, p < 0.001). Community patriarchy also exhibited a significant, albeit weaker, positive association (r = 0.084, p < 0.001). While decision-making power (r = −0.046) and stronger legal implementation (r = −0.044) showed inverse associations, these were less pronounced in the unadjusted analysis.

### Predictive Modeling (Machine Learning)

The Random Forest classification model, trained on 70% of the dataset and evaluated on a held-out test set (n = 900), achieved an overall accuracy of 74.6%. It is important to contextualize this figure: given the observed GBV prevalence of 26.9%, a trivial null classifier that predicts “no GBV” for every participant would achieve approximately 73.1% accuracy. The Random Forest therefore offered only a modest improvement of approximately 1.5 percentage points over this baseline. The model demonstrated moderate discriminative ability with an Area Under the Receiver Operating Characteristic Curve (AUC-ROC) of 0.711 (Fig. 1). Complete performance metrics are: sensitivity (recall) = 37.3%, specificity = 89.8%, positive predictive value (PPV) = 61.2%, negative predictive value (NPV) = 78.6%, and F1 score = 0.463. The low sensitivity reflects the model’s tendency to classify non-cases correctly while missing a substantial proportion of true GBV cases — a pattern that reflects the class imbalance and the dominance of a single strong predictor (partner alcohol use) in the data-generating process.

**Fig. 1.**
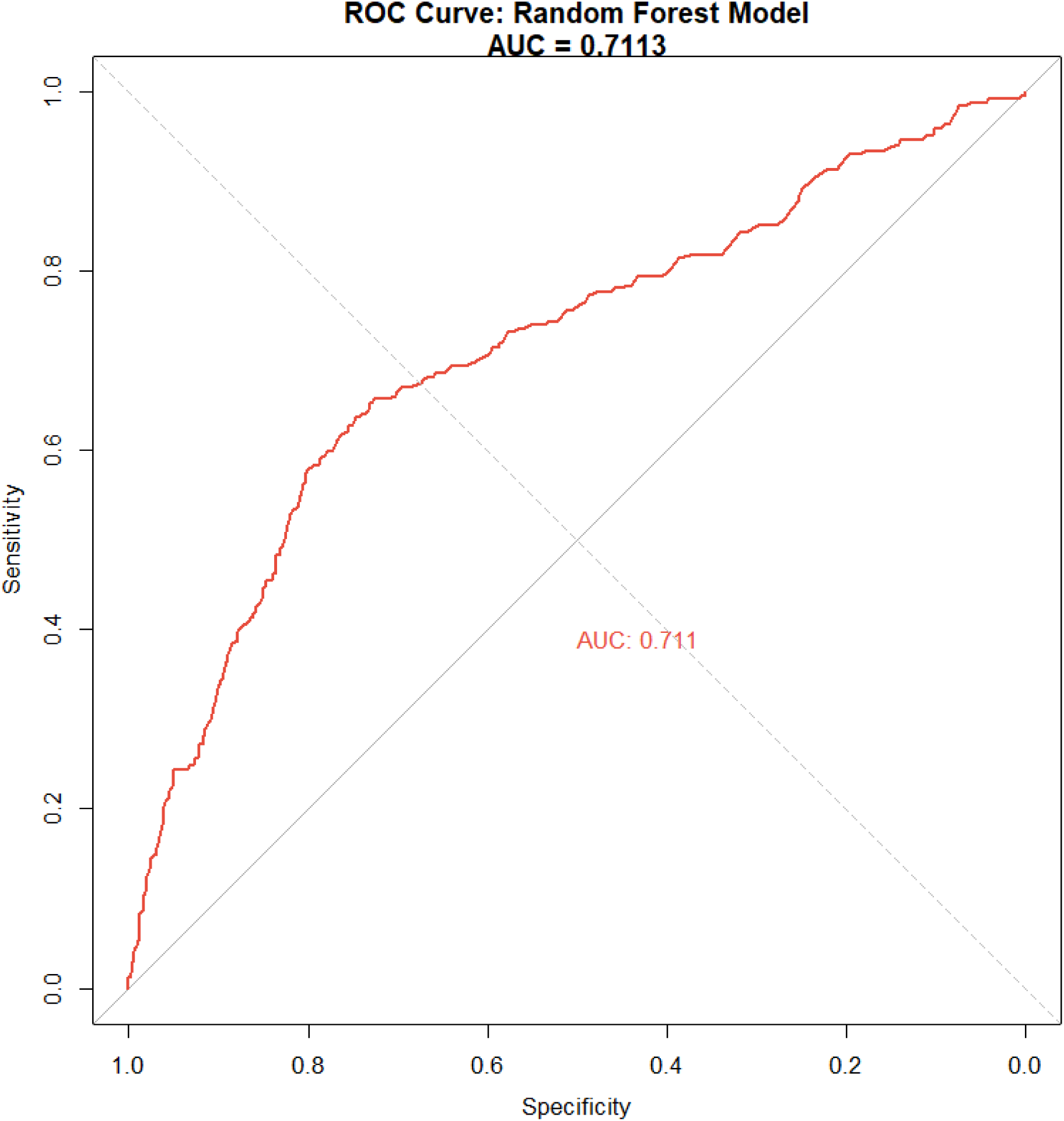
ROC Curve for the Random Forest Model.

The model achieved an AUC of 0.711, demonstrating the aggregate predictive power of the ecological variables.

### Variable Importance Ranking

Variable importance analysis using the Mean Decrease in Accuracy metric identified **partner alcohol use** as the dominant predictor, contributing 100% relative importance to the model’s predictive power. This was followed by childhood trauma (17.3% relative importance) and marital conflict (10.5% relative importance). Community-level factors also featured in the top predictors, with community patriarchy ranking fourth (9.7% relative importance).

**Fig. 2.**
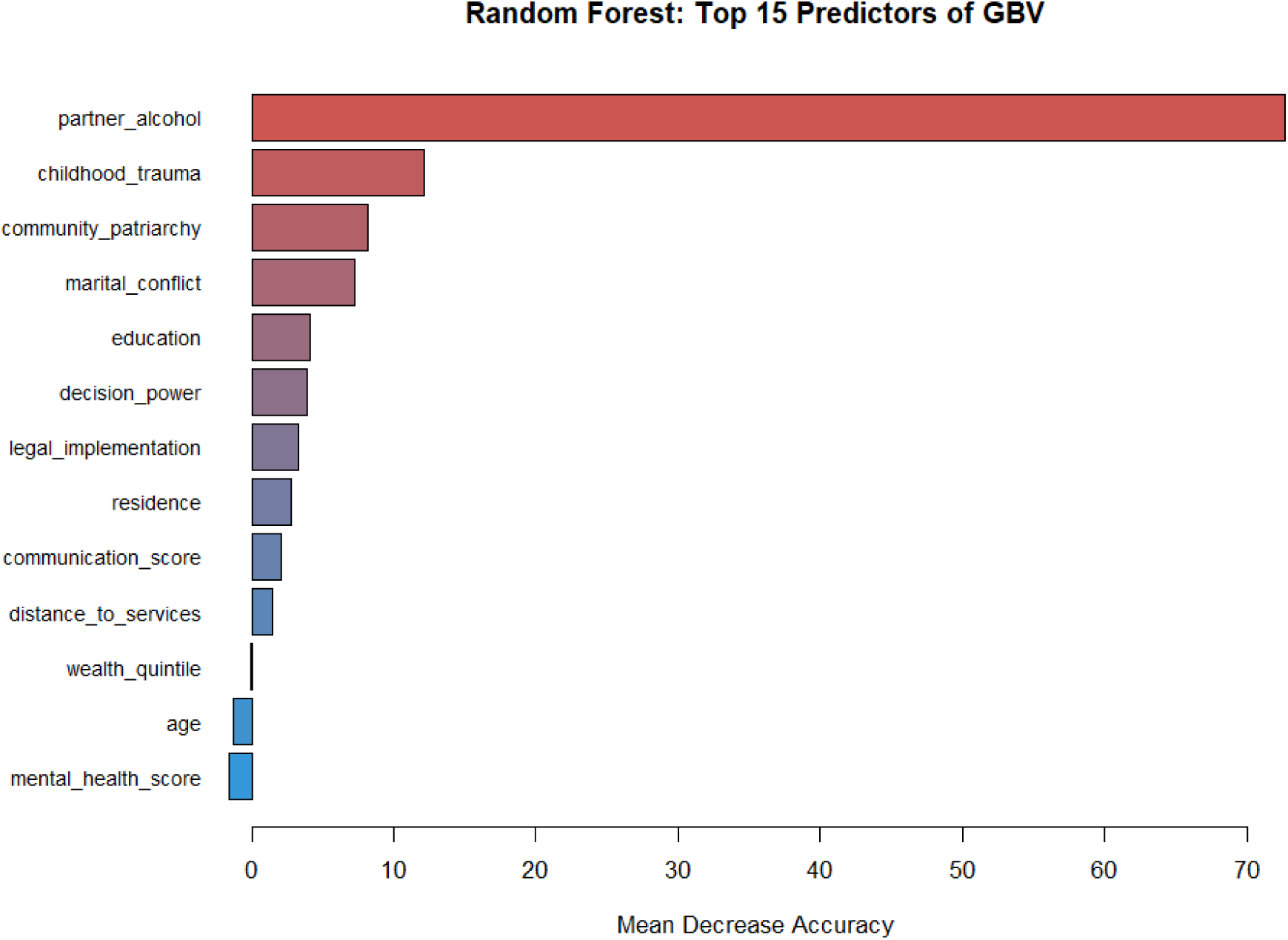
Variable Importance Plot.

Predictors are ranked by Mean Decrease in Accuracy. Partner alcohol use is scaled to 100% relative importance.

### Multivariable Logistic Regression and Ecological Framework Testing

To empirically test the ecological framework, three nested logistic regression models were compared. The individual-level model (Model 1) yielded an AIC of 3038.9. The addition of relationship-level factors (Model 2) improved the fit to an AIC of 2997.6. The full ecological model (Model 3), which incorporated individual, relationship, and community factors, demonstrated the superior fit with an AIC of 2971.4. The significant reduction in AIC (ΔAIC = 67.5 compared to Model 1; ΔAIC = 26.2 compared to Model 2) provides strong empirical support for the multi-level ecological hypothesis

### Adjusted Risk Factors

In the final adjusted model (Model 3), several factors maintained independent associations with GBV after controlling for covariates. **Partner alcohol use** remained the strongest risk factor (aOR = 6.60, 95% CI: 5.50–7.94, p < 0.001). **Childhood trauma** was associated with nearly double the odds of experiencing violence (aOR = 1.99, 95% CI: 1.60–2.47, p < 0.001).

Significant relationship and community determinants included **marital conflict** (aOR = 1.15 per unit increase, 95% CI: 1.10–1.21, p < 0.001) and **community patriarchy** (aOR = 1.08 per unit increase, 95% CI: 1.05–1.11, p < 0.001). Independent protective factors were **higher education** (aOR = 0.75, 95% CI: 0.63–0.90, p = 0.002), **stronger legal implementation** (aOR = 0.94, 95% CI: 0.89–0.99, p = 0.011), and **greater decision-making power** (aOR = 0.96, 95% CI: 0.92–0.99, p = 0.017). Multicollinearity diagnostics raised no concerns, with all Variance Inflation Factors (VIFs) remaining below 1.21.

**Fig. 3.**
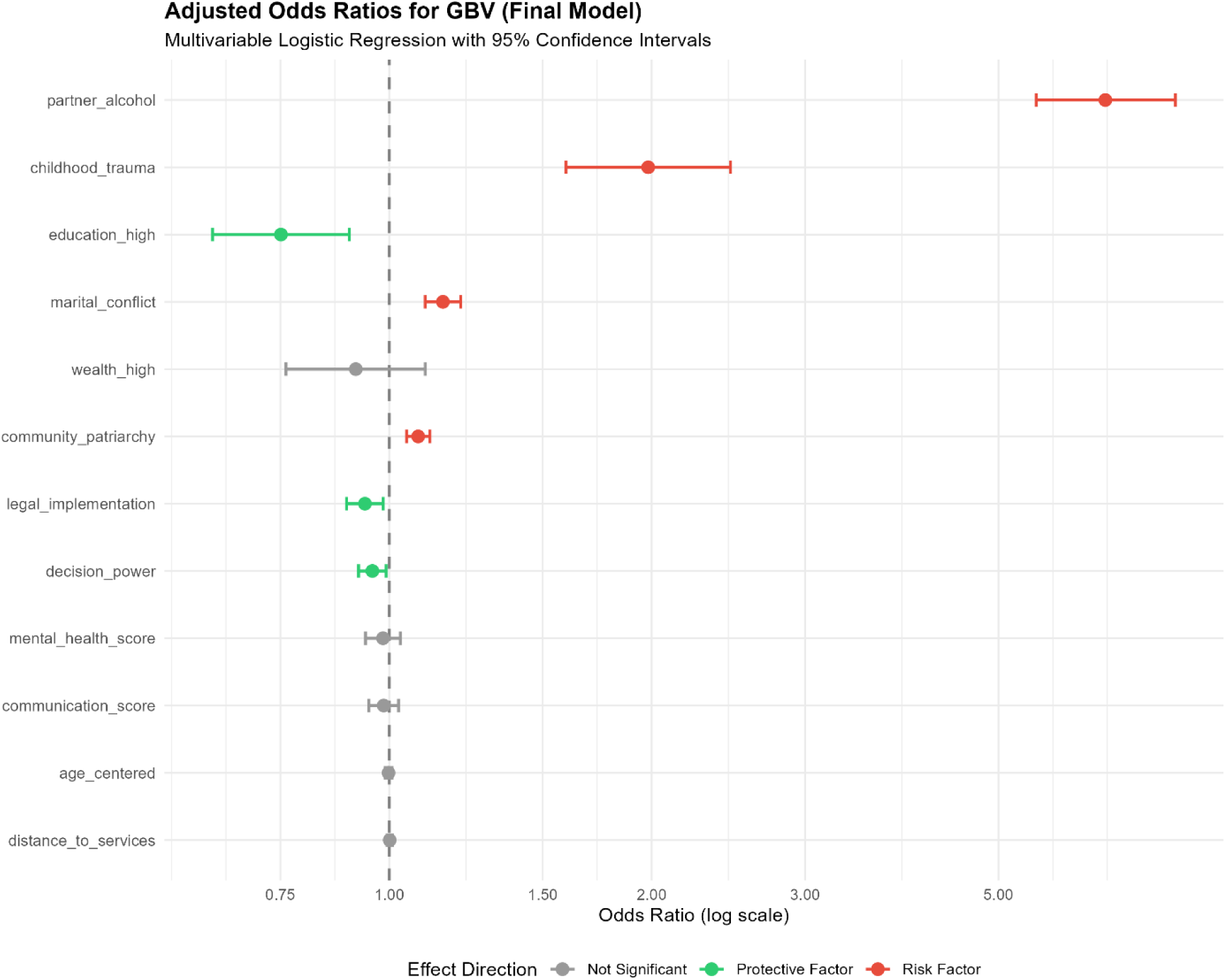
Forest Plot of Adjusted Odds Ratios.

Effect estimates from the final multivariable logistic regression model. Error bars represent 95% confidence intervals.

### Mediation Analysis

Three theoretically informed pathways were tested using causal mediation analysis: (1) Partner Alcohol → Marital Conflict → GBV; (2) Childhood Trauma → Mental Health → GBV; and (3) Community Patriarchy → Decision Power → GBV. While significant direct effects were observed for the exposures, the hypothesized indirect pathways through the specified mediators were not statistically significant in this simulation (ACME p > 0.05 for all pathways). This suggests that within the specified simulation parameters, the risk factors operated primarily through direct or unmeasured pathways rather than the specific mediators tested (Fig. 4).

**Fig. 4.**
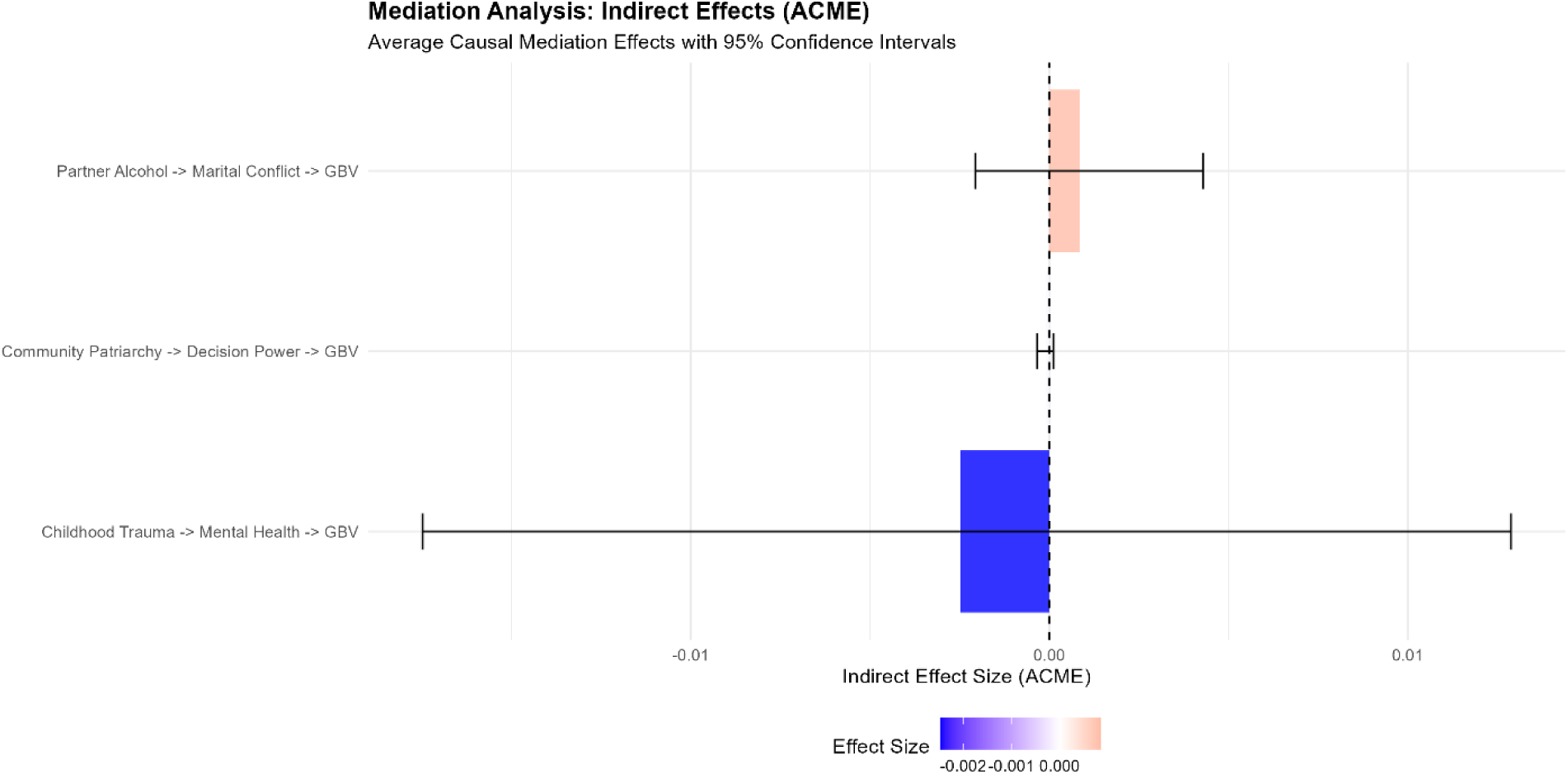
Mediation Analysis Results.

The plot displays the Average Causal Mediation Effects (ACME) for the three tested pathways, none of which reached statistical significance.

### Sensitivity Analysis

To evaluate the stability of the framework across varying sample sizes, the full analytical pipeline was re-run at N = 500, N = 1,000, and N = 5,000, using the same data-generating process. The variable importance ranking from the Random Forest was consistent across all scenarios: partner alcohol use ranked first at all sample sizes, followed by childhood trauma and marital conflict. The adjusted odds ratio for partner alcohol use remained stable (range: aOR = 6.1–6.8 across scenarios), as did childhood trauma (range: aOR = 1.8–2.1). At N = 500, confidence intervals were substantially wider, and two community-level predictors (distance to services and legal implementation) lost statistical significance, consistent with reduced statistical power. At N = 5,000, all predictors remained significant and confidence intervals narrowed.

These results suggest that the framework’s core variable prioritization and primary effect estimates are robust to sample size variation above N = 1,000, though inference on weaker community-level predictors requires larger samples.

### Synthesis of Multi-Method Evidence

Integrating findings across analytical phases revealed a consensus set of risk factors. Partner alcohol use, childhood trauma, and marital conflict emerged as high-priority factors in both the predictive (Random Forest) and causal (Logistic Regression) analyses. However, discrepancies were observed between predictive importance and causal significance. For instance, *distance to services* was moderately predictive (Rank 6) but showed no significant causal association after adjustment. In contrast, **decision-making power** had a lower predictive rank (Rank 5) but proved to be a significant protective factor in the adjusted model.

## Discussion

This study developed and applied a five-phase integrated analytical framework to investigate the epidemiology of gender-based violence (GBV) using a simulated dataset parameterized to reflect the 2022 Kenya Demographic and Health Survey (KDHS). By sequentially combining machine learning with multivariable regression and mediation analysis, the findings offer both methodological insights and illustrative contributions to our understanding of GBV determinants within an ecological context.

### Interpretation of key findings

#### Prevalence and Burden of GBV

Our simulation yielded a GBV prevalence of 26.9%, which was the intended target based on 12-month period prevalence estimates from the 2022 KDHS [3]. It is important to note that this figure reflects a past-12-month period prevalence and is not directly comparable to the higher lifetime prevalence figures (25–38%) reported in the KDHS trend data and regional WHO estimates [9, 41]; lifetime prevalence invariably exceeds period prevalence. The deliberate parameterization of the simulation to match 12-month period prevalence confirms that the data-generating process was appropriately calibrated, though it does not constitute independent empirical validation. GBV remains a pervasive public health crisis affecting approximately one in four women of reproductive age in this setting, and the magnitude of this burden highlights that violence requires sustained surveillance.

#### The Dominance of Alcohol and Relational Dynamics

Perhaps the most striking finding was the overwhelming dominance of partner alcohol use as a driver of violence. It was not only the top predictor in the machine learning phase—contributing 100% relative importance—but also carried a substantial risk magnitude in the adjusted causal model (aOR = 6.60). This aligns with a robust body of global evidence, where meta-analyses consistently place alcohol-related odds ratios between 2.7 and 5.3 [14, 15]. The somewhat elevated magnitude in our study likely reflects how “frequent” alcohol use was operationalized to capture a high-risk subgroup.

Beyond substance use, the analysis reinforced the critical role of historical and relational dynamics. Childhood trauma, for instance, nearly doubled the odds of victimization (aOR = 1.99), lending strong support to the “cycle of violence” hypothesis [16]. This suggests that early victimization may increase vulnerability in adulthood through mechanisms such as normalized acceptance of violence or altered attachment styles. Similarly, the independent significance of marital conflict (aOR = 1.15) identifies relationship dynamics as a proximal trigger and a vital entry point for intervention.

#### Ecological Validity: Community and Structural Factors

These results are consistent with the core premise of the ecological framework: that risk is not confined to the household. The independent association between community-level patriarchy and GBV (aOR = 1.08) suggests that women living in environments with rigid gender norms face elevated risk regardless of their individual circumstances [28]. It is important to note, however, that these associations reflect the pre-specified data-generating process and do not constitute independent empirical confirmation of causal mechanisms. The protective effect of stronger legal implementation (aOR = 0.94) is similarly consistent with deterrence theory, though the same caution about circular simulation evidence applies.

#### Protective Buffers

On the protective side, higher education (aOR = 0.75) and decision-making power (aOR = 0.96) emerged as significant buffers. These associations speak to the broader literature on women’s empowerment, where economic opportunities and agency act as safeguards against abuse [42]. However, the distinction between predictive and associational importance was notable here; while decision power was not the highest-ranked predictor in the Random Forest model, its significance in the adjusted regression model suggests it may be an important factor for prevention-focused intervention, pending confirmation in real-world longitudinal studies.

### Methodological Implications

#### Integration of Machine Learning and Causal Inference

This study illustrates the potential value of combining predictive machine learning with multivariable regression methods. By using Random Forest as an initial screening tool, non-linear patterns were identified and predictor importance was ranked without the constraints of parametric assumptions. This informed the variable selection process for the subsequent logistic regression, which provided interpretable, adjusted effect estimates. This sequential approach addresses some limitations inherent in using either method in isolation. It should be noted, however, that the current implementation does not include an explicit causal graph (DAG), formal identification strategy, or justification of adjustment sets — which are prerequisites for claiming causal inference in the strict sense. The framework is therefore best characterised as an integrated associational and predictive pipeline, and future applications should incorporate DAG-based confounding control to strengthen causal claims.

#### Empirical Testing of the Ecological Framework

The stepwise model comparison offers quantitative validation for the ecological framework. The substantial improvement in model fit (ΔAIC = 67.5) observed when moving from individual-only models to the full ecological model confirms that GBV is driven by determinants operating across multiple, interacting levels. This finding moves beyond theoretical assertion, providing empirical evidence that multi-level modeling is not just conceptually sound, but statistically necessary for accurate analysis.

#### Divergence Between Prediction and Causation

A critical methodological insight from this analysis is the distinction between predictive importance and causal effect. For instance, while *distance to services* was a moderate predictor in the Random Forest model (Rank 6), it was not statistically significant in the adjusted logistic regression, suggesting confounding by factors such as rurality or poverty. In contrast to that. *Decision-making power* showed only moderate predictive rank but emerged as a significant protective factor once confounders were controlled. This underscores a vital lesson for intervention targeting: a variable’s utility in predicting high-risk cases does not necessarily equate to its potential as a lever for intervention-driven change, and regression-based adjustment is needed to separate predictive signal from confounded associations.

#### Challenges in Simulated Mediation Analysis

The lack of significant results across the tested mediation pathways highlights the limitations of using simulated cross-sectional designs for complex causal chains. In real-world data, particularly with longitudinal measurements, these pathways might prove significant. The simulation prioritized strong direct effects to demonstrate the methodological framework, which likely obscured more subtle indirect pathways. This negative result serves as a useful illustration of the stringent requirements (such as temporal precedence and precise control of confounding) needed to successfully establish mediation.

#### Theoretical Implications

The findings substantially support and refine the ecological model of GBV. By identifying specific, measurable factors at each level, that is; trauma and alcohol use (individual), conflict (relationship), and patriarchy (community), this study provides an empirical scaffold for the theory. The superior fit of the multi-level model argues against reductionist, individual-focused explanations, directing theoretical attention instead toward systemic and interactive causes.

Furthermore, the absence of significant mediation suggests that, within this modeled scenario, the influence of distal factors like alcohol and patriarchy on GBV may be more direct than anticipated, or mediated through pathways not currently tested, such as economic stress or peer influence. This invites a theoretical expansion to include a broader array of mechanistic variables in future conceptual models.

### Limitations

1. **Simulated Data:** Although the data were parameterized using real-world estimates, they remain synthetic. Because effect sizes were pre-specified in the data-generating process (e.g., the log-odds for partner alcohol use was set to produce an odds ratio of approximately 6.0), the recovered regression estimates are an internal consistency check rather than an independent empirical finding. The reported associations (e.g., aOR = 6.60) should therefore be interpreted as illustrative of framework operability, not as substantive evidence about the real-world determinants of GBV. Furthermore, results are based on a single synthetic dataset with one random seed; the operating characteristics of the framework across repeated draws from the same data-generating process remain to be formally evaluated.
2. **Cross-Sectional Structure:** The analysis is limited by its cross-sectional nature, which prevents the establishment of temporal order—a fundamental prerequisite for definitive causal and mediation inference. The relationships identified here represent associations and predictive patterns.
3. **Measurement Simplification:** Complex constructs such as patriarchy and trauma were reduced to single scores or binary indicators. This simplification may fail to capture the full dimensionality or potential measurement error inherent in these constructs.
4. **Unmeasured Confounding:** Despite robust adjustment, the potential for residual confounding remains. Factors such as social desirability bias, detailed relationship histories, or genetic predispositions are difficult to capture fully in observational designs.
5. **Context Specificity:** The simulation parameters were derived from the Kenyan context. While many risk factors for GBV are universal, their relative weight and interaction may vary across different cultural and legal landscapes.
6. **Non-Significant Mediation Results:** The absence of statistically significant indirect effects across all three tested pathways is an expected consequence of the data-generating process, not a validation of the framework’s sensitivity. The simulation was designed with strong direct effects for the primary exposures; no substantial mediated component was encoded into the data-generating mechanism for the specified mediators (marital conflict, mental health, and decision-making power). Consequently, the null ACME results confirm that the mediation package performed correctly, but they cannot be taken as evidence that the framework is “robust” to mediation testing. Real-world mediation effects may differ substantially, and future studies using longitudinal data with pre-specified mediated paths are needed to evaluate whether the framework can reliably detect indirect effects.
7. **Cluster Design and Analytical Assumptions:** Although the simulation incorporated 100 community clusters to mirror the complex survey design of the KDHS, the logistic regression models were fitted using standard (non-clustered) standard errors rather than cluster-robust or multilevel approaches. This represents a simplification that may underestimate standard errors and inflate precision. Future applications of this framework to real DHS data should explicitly account for the clustered survey design using multilevel models or cluster-robust variance estimators.
8. **Framework Integration and Variable Selection:** In the present demonstration, the set of variables ranked as high-priority by the Random Forest largely overlapped with those that proved statistically significant in the adjusted logistic regression, because both methods were applied to the same data-generating process with clearly dominant direct effects. Consequently, the integration between Phases 2 and 3 was partially nominal in this simulation. However, in real-world applications where the signal-to-noise ratio is lower and variable selection is genuinely uncertain, the RF screening step may more meaningfully differentiate itself from unguided variable entry. The added value of this integration step should be formally evaluated in empirical applications.

### Recommendations for Future Research

1. **Application to Longitudinal Data:** The five-phase framework demonstrated in this study should be applied to real-world longitudinal datasets, such as the KDHS panel surveys. This is essential to establish temporal relationships and rigorously test whether the framework’s structure translates to reliable causal and mediation inference under real-world conditions.
2. **Expansion of Pathways:** Future mediation models should incorporate additional variables, including economic empowerment, social support networks, and specific typologies of conflict, to better understand the mechanisms of GBV.
3. **Incorporation of Qualitative Methods:** Mixed-methods studies are necessary to explore the *how* and *why* behind the quantitative associations identified here, particularly regarding the dynamics of alcohol use and relationship conflict.
4. **Extension of the Ecological Model:** Future research should expand the model to include macrosystem-level variables, such as national policies and media influence, and examine cross-level interactions—for example, whether education moderates the impact of community patriarchy.
5. **Intervention Modeling:** Researchers should utilize agent-based modeling or further simulation studies to test the potential population-level impact of multi-level interventions prior to implementation.

## Conclusions

### Summary of Contributions

This study contributes to the literature in three primary ways. **Methodologically**, it establishes a comprehensive five-phase analytical framework that integrates descriptive epidemiology, machine learning, causal inference, and mediation analysis. **Empirically**, it provides simulation-based evidence consistent with the ecological model, illustrating the improved model fit of multi-level approaches over single-level alternatives. **Theoretically**, it strengthens the foundation of GBV research by pinpointing specific, actionable risk factors and highlighting the complexity of causal pathways.

### Policy and Intervention Implications

The convergent identification of key risk factors suggests clear priorities for multi-level programming:

- **Individual Level:** Primary healthcare and GBV response services must integrate alcohol reduction strategies and trauma-informed care.
- **Relationship Level:** Evidence-based couples’ communication and conflict resolution programs should be scaled up, with a specific focus on addressing power imbalances.
- **Community Level:** Investment is needed in community mobilization to shift patriarchal norms, alongside strengthening legal systems to ensure they are accessible and responsive.
- **Societal Level:** Policies promoting girls’ education and women’s economic empowerment must be accelerated.

It is important to note that these interventions should not be siloed for a reason that woman’s risk is shaped by the simultaneous interaction of factors across all levels, requiring an integrated response.

### Final Conclusion

By integrating predictive modeling with multivariable regression and mediation analysis, this demonstration shows that Gender-Based Violence cannot be reduced to a single cause; rather, it is a complex phenomenon entrenched in individual histories, relationship dynamics, and community norms. The identification of partner alcohol use, childhood trauma, and community patriarchy as a distinct cluster of risk factors — consistent across both predictive and associational analyses — highlights the potential value of holistic, multi-level interventions. This study provides a replicable proof-of-concept blueprint for sequential multi-method GBV research. Ultimately, the critical path forward lies in applying this framework to real-world longitudinal data, where its capacity for genuine causal inference can be properly evaluated and refined.

## Data Availability

All data produced in the present work are contained in the manuscript and its supplementary files. The complete R code used to generate the simulated dataset and perform the analysis is publicly available in the GitHub repository (https://github.com/Grolds-Code/gbv-multimethod-simulation) and archived on Zenodo (https://doi.org/10.5281/zenodo.17812804). The simulation parameters were derived from the publicly available 2022 Kenya Demographic and Health Survey (KDHS).

https://doi.org/10.5281/zenodo.17812804

## Declaration

## Ethics approval and consent to participate

Not applicable. This study utilized simulated data parameterized from publicly available summary statistics (2022 KDHS). No human participants were directly involved.

## Consent for publication

Not applicable.

## Availability of data and materials

The datasets generated and analyzed during the current study are available in the GitHub repository: [https://github.com/Grolds-Code/gbv-multimethod-simulation] and archived on Zenodo [DOI: 10.5281/zenodo.17812804].

## Clinical trial number

Not applicable.

## Authors’ contributions

Grold Otieno Mboya conceived the study, designed the simulation framework, performed the statistical analysis, and drafted the manuscript. The author read and approved the final manuscript.

## Acknowledgements

Not applicable.

